# Anxiety and depressive symptoms in women with disabilities during pregnancy and childbirth: an analysis of Nepal demographic and health survey data

**DOI:** 10.1101/2025.03.18.25324168

**Authors:** Hridaya Raj Devkota, Pratik Adhikary, Jaslina Bohora, Kalyan Lama, Sasmita Poudel

## Abstract

**Background:** Evidence reveals an association between depressive symptoms and disability, while increased incidences of anxiety and depression are observed among women during pregnancy and childbirth. However, the experience of anxiety and depressive symptoms among women with disabilities is rarely studied in Nepal.

**Objective:** Determine the prevalence and factors associated with anxiety and depressive symptoms among reproductive-age women during pregnancy and delivery and compare their magnitude and severity between those with and without disabilities.

**Methods:** Data from the Nepal Demographic and Health Survey - 2022, was analyzed. The survey used the Patient Health Questionnaire (PHQ-9) and Generalized Anxiety Disorder (GAD-7) scales for depression and anxiety measurements, respectively. ANOVA was used to compare data with multiple means across different groups, and logistic regression explored associations between outcomes and independent variables.

**Results:** The overall prevalence of depression and anxiety among women aged 15 - 49 years was found at 5.9% and 22.2%, respectively. Most (78.4%) did not have depressive symptoms, while 15.7% had mild, 4.2% had moderate, and only 1.7% had severe depressive symptoms. Regarding anxiety, 77.8% had minimal or no symptoms, 20.8% had moderate symptoms, and only 1.4% had severe anxiety symptoms. Women with disabilities were 1.72 times more likely to experience depressive symptoms (OR 1.72, 95% CI 1.51 – 1.97; P<0.001), and 1.8 times more likely to experience anxiety (OR 1.80, 95% CI 1.56 – 2.05; P<0.001) compared to women without disabilities. Marital status, caste, and ethnicity were strongly associated with depression and anxiety (P<0.001), while pregnancy, religion, and household wealth were associated with depression (P<0.05).

**Conclusion:** This study highlights the substantial mental health challenges among women of reproductive-age in Nepal, particularly those with disabilities, during pregnancy and childbirth. The study urges the prompt implementation of focused interventions and policies to address these disparities and improve the well-being of reproductive-age women in Nepal, with special attention to those with disabilities.

## Introduction

An estimated 4.4% of the world’s population suffers from major depressive disorders, and a comparable percentage suffers from anxiety disorders accompanied by depressive symptoms [1]. According to the findings of the Global Burden of Disease 2019 assessment, depression ranked as the second most prevalent contributor to the years lived with disability, while anxiety disorders occupied the eighth position among the top 25 causes in all age groups. People with disabilities are frequently more vulnerable to mental health problems. Substantial evidence shows that people with physical disabilities are three times more likely to experience depression compared to their non-disabled counterparts [2–4]. Those in this category, as opposed to those without disabilities, frequently encounter challenging living conditions and require specialized support, contributing to increased poverty rates and weakened social connections [5]. In particular, the age-standardized rate of disability adjusted life years attributed to mental disorders is higher among women than among men. Furthermore, certain groups, such as pregnant women, tend to be more vulnerable to anxiety and depressive symptoms due to the multiple transitions that comprise roles, psychological dynamics, and physiological changes [6].

Mental health conditions are common complications of pregnancy and also an underlying cause of suicide and pregnancy-related deaths. The onset of depressive episodes after delivery develops at a critical moment in a woman’s life and can continue for long periods [7,8]. The probability of depressive episodes throughout pregnancy can be twice as high as during other periods of a woman’s life [9]. Furthermore, women with disabilities are at increased risk of experiencing symptoms of depression than other women without disabilities after giving birth [10]. The exact prevalence rates of depression and anxiety can vary depending on factors such as type of disability, cultural differences, socioeconomic conditions, and access to healthcare. A systematic review study suggests that the global prevalence of depression among postpartum women is 17.22%, ranging from 6.48% in Denmark to 60.93% in Afghanistan. The same study reported the prevalence rate of postpartum depression prevalence in Nepal at 16.41% [11].

Depression and anxiety constitute the predominant mental health disorders in women in both low and high income countries; however, women in low income countries are more at high risk [12,13]. Risk factors for depression in pregnant women in low and middle-income countries included physical, psychological or sexual abuse in early childhood or adulthood, limited maternal education, lower socioeconomic status, and insufficient social support [14,15]. A qualitative study in Vietnam suggested that Vietnamese women with physical disabilities experience great anxiety during pregnancy [16]. A study in South Africa revealed that women in a low-income group are at higher risk of perinatal depressive symptoms [17]. Furthermore, women of lower family income in Kuwait were reported to be at increased risk of depressive symptoms during pregnancy [18]. In Malaysia, inadequate social support poses a risk factor for experiencing depressive symptoms during the antenatal period [19]. Similar to other Asian countries, low level of education, lower social support from spouses, family, and friends, poor living conditions, severe food insecurity, perinatal health problems, poor antenatal care, and sex preference (son) or sex of the baby were commonly reported risk factors for anxiety and depression among women during pregnancy in Nepal [20,21].

Women in low- and middle-income countries (LMICs) are believed to carry a disproportionate burden of mental health disorders. However, the prevalence and associated factors of these disorders during pregnancy among women with disabilities have not been extensively studied. The current understanding of the epidemiology of anxiety and depression is based mainly on a few geographic surveys and very limited national evidence. The escalation rates of depression and anxiety among pregnant women and birthing women with disabilities in Nepal have become a growing health problem. However, there remains a dearth of scholarly research on the determinants associated with depression among women with disabilities in Nepal. We hypothesized that people with disabilities are more likely to experience anxiety and depressive symptoms. This study aimed to explore the occurrence and factors that influence anxiety and depression in women with disabilities during pregnancy and after childbirth, using nationally representative data.

## Methodology

### Data Source

The study analyzed secondary data from the Nepal Demographic and Health Survey (NDHS) 2022, which was conducted by New ERA under the direction of Nepal’s Ministry of Health and Population (MoHP) with technical support from the International Classification of Functioning (ICF). USAID funded the study. NDHS 2022 provides current information on key demographic and health indicators in Nepal. This was the sixth periodic nationally representative survey conducted since 1996, and, notably, it was the first to include the mental health and disability module.

### Study Population and Sample

In this analysis, all women aged 15 to 49 years were considered the study population. A total sample of 7442 women in this age group who participated in the disability and mental health module questionnaires were extracted from the NDHS data sets and analyzed.

NDHS was the national survey and all households in the country were the universe of the study. The survey used a two-stage stratified sampling design to make the results representative. The sampling frame used was an updated version of the Nepal Population and Housing Census 2011 frame. Dividing the seven provinces into urban and rural, 14 sampling strata were created. In the first stage, considering the districts of the 2011 census as sub-districts, 476 primary sampling units (PSU) or enumeration areas (EA) were selected in urban (248) and rural (228) areas. In the second stage, a random sample of 30 residential dwelling units (DU) was selected from each PSU for a total sample of 14,280 households. All men and women aged 15 to 49 years who were permanent residents of the selected households were recruited for interviews. Detailed information on survey design, sampling methods, survey procedure, recruitment, and response rates is available in the NDHS final report [22].

### Survey instrument and data collection procedure

NDHS 2022 used six different sets of standard questionnaires that included various module questions to collect individual and household-related information. The questionnaires included questions for background and sociodemographic information about the home and family members, the home and possessions of the home, the assets and ownership, disability and mental health status. The Washington Group Disability Statistics Short Set on Functioning (WG-SS) questionnaire was used to assess disability status. The questionnaire included six core functional domains: seeing, hearing, communication, cognition, walking, and self-care. The severity scale of the full spectrum of functioning, from mild to severe, was used [23]. Similarly, the ‘General Anxiety Disorder’ (GAD-7) and the ‘Patient Health Questionnaire’ (PHQ-9), developed by the Drs. Robert L. Spitzer, Janet B.W. Williams, Kurt Kroenke, and colleagues (1999) were used to collect and measure anxiety and depression symptoms data [24]. Both PHQ-9 and GAD-7 are self-report scales consisting of nine and seven items, respectively. The PHQ-9 measures the frequency of depression symptoms (eg feeling hopeless, having little interest or pleasure in doing things, and negative self-evaluation), while the GAD-7 scale measures the frequency of generalized anxiety symptoms (eg feeling nervous, not being able to stop or control worrying, becoming easily annoyed or irritable).

The questionnaires were translated into three languages, Nepali, Maithili, and Bhojpuri, and pre-tested before field administration. Well-trained field researchers collected data between January and June 2022. All of this data was cleaned, primarily coded, and available in the public domain. Data were extracted from the set of household questionnaires for analysis.

### Measure

The outcome variable(s) of this study were mental health status, that is, depression and anxiety. The prevalence of anxiety and depressive symptoms and their severity were evaluated and compared between i) women with and without disabilities, ii) pregnant and nonpregnant women, and iii) women who had given birth within the past year were evaluated and compared to those who had not. The scores generated using the PHQ-9 scale range from 0 to 27. We consider a score of 0 – 4 ‘minimal or no symptoms’, 5 – 9 ‘mild’, 10 - 14 ‘moderate’, 15 – 19 ‘moderately severe’ and 20 – 27 ‘severe’. [24]. The GAD-7 score ranges from 0 to 21, and is considered a score of 0 - 5 ‘mild or no anxiety’, 6 – 14 ‘moderate’, and 15 – 21 ‘severe’ [25].

Disability and severity scale based on the WHO-defined International Classification of Functioning (ICF) on Disability and Health assessed using the Washington Group questionnaire (short set), current pregnancy and birth in the past year were used as covariates, and sociodemographic variables such as place of residence, age, caste and ethnicity, religion, education, occupation, marital status, and household wealth index were independent variables.

Table 1 presents the key variables used in the study and their definitions.

**Table 1:** Variables and their definitions used in the study

| Variables | Definition |
| --- | --- |
| **Outcome Variables** |  |
| i) Depression | PHQ-9 scale score less than five was considered no depression, and five or above was considered depression. |
| ii) Anxiety | GAD-7 scale score less than five was considered no anxiety, and five or more was considered anxiety. |
| **Covariate** |  |
| i) Disability | As defined by WHO's ICF, six functional domains were included in the analyzes - seeing, hearing, communicating, remembering or concentrating, walking or climbing steps, and washing or dressing.<br><u>Severity scale:</u> The level of activity limitation or difficulty is classified as no difficulty, some difficulty, a lot of difficulty, and cannot do at all. |
| ii) Pregnancy | Respondents reported pregnancy at the time of the survey. |
| iii) Birth past year | Birth given by the respondent within one year |
| **Sociodemographic variables** |  |
| Place of residence | Respondent's place of residence: rural or urban. |
| Age | Completed age in years of respondent at the time of survey |
| Caste and ethnicity | Self-reported caste and ethnic group of the respondents classified as Brahmin / Chhetri, Jana Jaati, Dalit, Tarai caste (Madhesi, Muslim, and others) |
| Religion | Respondent's self-reported religious belief at the time of survey |
| Education | Respondent's reported level of educational attainment - no education, basic level education, secondary and higher-level |
| Occupation | Respondent's reported main occupation at the time of the survey categorized as professional/technical/managerial, clerical, sales and services, skilled manual, unskilled manual, agriculture, and other. |
| Marital status | Respondent's self-reported status of the respondent at the time of the survey was classified as never married, married or living together with a partner at the time of the survey, or single (widowed, divorced or separated). |
| Household wealth index | Scores based on the number and kinds of consumer goods that households own, ranging from a television to a bicycle or car, and housing characteristics such as source of drinking water, toilet facilities, and flooring materials. These scores are derived using the principal component analysis. The household population is ranked by the score, which is divided into five equal categories (20% each). |

### Statistical analysis

The extracted data were transferred to IBM SPSS statistics version 23 for further cleaning, recoding, and analysis. The weighted sample from the survey was used. We use descriptive and inferential statistics. The categorical variables were summarized using frequency and percentage and the continuous variables using mean and standard deviation (SD). We applied one-way analysis of variance (ANOVA) to compare data with multiple means in different groups and carried out logistic regressions to investigate the association between outcome and independent variables and estimate the odds ratio between them with 95% confidence intervals. A p-value of <0.05 was considered statistically significant.

### Ethical Considerations

The NDHS 2022 survey protocol was reviewed and approved by the Nepal Health Research Council, Kathmandu (Ref. No. 678, 30 September 2021) and the ICF Institutional Review Board, Rockville, Maryland, USA (Ref. No. 180657.0.001.NP.DHS, 28 April 2022). However, the study was guided by secondary data usage standards, such as maintaining deidentified data, ensuring that the results of the analysis do not reidentify participants, and using the data do not cause any damage or distress to the participants.

## Results

The results of this study are based on the analysis of a total of 7442 women aged 15 to 49 years to whom the questionnaires for the PHQ-9, GAD-7, and disability module were administered.

### Characteristics of study samples

Of the total sample, just under 8% of women gave birth in the last year and nearly 4% were pregnant at the time of the survey. Approximately 23% of respondents reported having’some difficulty,’ 1.8% reported having ‘a lot of difficulty,’ and a few (0.2%) said ‘cannot do at all.’ The mean age of the study women was 29.8 (SD ±9.8) years, and more than half (54%) were urban residents. Most of the respondents (84.7%) reported their religious belief as Hinduism. Approximately an equal proportion of the participants (6% each) reported that they believe in Buddhism and Christianity, while Muslims were 3.7%. The survey included a significant proportion of Jana Jaati ethnic groups (indigenous) (34.7%), followed by Brahmin and Chhetries (33.2%). More than one in six women were Dalits, while participants in Madhesi, Muslims, and other Tarai castes and ethnic groups together were 15.4%. Brahmin and Chhetri are considered the highest and most privileged groups, and Dalits are considered the lowest in the caste hierarchy and historically excluded and marginalized groups. Almost one in five women said they were never married, and a small portion of the sample (3.2%) reported their current marital status as single, widowed, divorced or separated. More than a quarter (27.3%) of the study participants had no school education and less than 3% reported higher education. Just under one-fourth of the study participants reported not working, while approximately 63% reported that their main occupation was manual work. More than 6% of the respondents reported that their main occupation was a professional or service job, and about 7% said sales and services were their main occupation. More than 27% of the sample participants belonged to the poorest household wealth quintile, while 13.8% belonged to the richest wealth quintile. (Table 3)

### Prevalence of depression and anxiety

The overall mean scores for PHQ-9 and GAD-7 were 2.7 (SD ±3.7) and 3.4 (SD ±3.6), respectively. Of the total, 78.4% of the participants did not have depressive symptoms, 15.7% had mild symptoms, 4.2% had moderate symptoms, and 1.7% had severe depressive symptoms. Similarly, 77.8% had minimal or no anxiety, 20.8% had moderate anxiety, and 1.4% showed severe anxiety symptoms.

Using a cut-off point of 10 for the PHQ-9 score and 6 for the GAD-7 score, the analysis detected the prevalence of depression at 5.9% and the prevalence of anxiety at 22.2% among 15 - 49 year old women. The prevalence of depression and anxiety in women with disabilities was significantly higher than in women without disabilities (Depression: 9.3% vs. 5%, P<0.001; Anxiety: 32% vs. 19.4%, P<0.001). However, there were no significant differences in The prevalence of depression and anxiety prevalence between pregnant and non-pregnant women, and between women who gave birth in the past year and those who did not. (Table 2).

**Table 2:** Distribution of depression and anxiety score by disability, pregnancy, and birth within one year

| Factors | Depression Grading (n= 7442) |  |  |  |  | Anxiety Grading (n=7442) |  |  |  |
| --- | --- | --- | --- | --- | --- | --- | --- | --- | --- |
|  | 0 - 4 | 5 - 9 | 10 - 14 | 15 - 27 | P value | 0 - 5 | 6 - 14 | 15 - 21 | P value |
| **All** | 5833 (78.4%) | 1167 (15.7%) | 315 (4.2%) | 127 (1.7%) | - | 5790 (77.8%) | 1546 (20.8%) | 106 (1.4%) | - |
| **Disability** |  |  |  |  |  |  |  |  |  |
| No | 4668 (80.9%) | 814 (14.1%) | 202 (3.5%) | 84 (1.5%) | **0.000** | 4646 (80.6%) | 1052 (18.3%) | 63 (1.1%) | **0.000** |
| Yes | 1172 (69.7%) | 353 (21.0%) | 113 (6.7%) | 43 (2.6%) |  | 1144 (68.1%) | 494 (29.4%) | 43 (2.6%) |  |
| **Pregnancy** |  |  |  |  |  |  |  |  |  |
| No | 5627 (78.6%) | 1112 (15.5%) | 298 (4.2%) | 120 (1.7%) | 0.073 | 5568 (77.8%) | 1488 (20.8%) | 101 (1.4%) | 0.881 |
| Yes | 206 (72.3%) | 55 (19.3%) | 17 (6.0%) | 7 (2.5%) |  | 222 (77.9%) | 58 (20.4%) | 5 (1.8%) |  |
| **Birth in past year** |  |  |  |  |  |  |  |  |  |
| No | 5374 (78.2%) | 1081 (15.7%) | 292 (4.3%) | 121 (1.8%) | 0.560 | 5341 (77.8%) | 1427 (20.8%) | 100 (1.5%) | 0.725 |
| Yes | 459 (80.0%) | 86 (15.0%) | 23 (4.0%) | 6 (1.0%) |  | 449 (78.2%) | 119 (20.7%) | 6 (1.0%) |  |

**Depression:** 0 – 4 minimal symptoms or no depression, 5 – 9 mild, 10 – 14 moderate, 15 – 27 severe. (Kroenke et al. 2001); **Anxiety:** 0 – 5 mild, 6 – 14 moderate, 15 - 21 severe. (Spitzer et al. 2006b). For comparability purposes, a PHQ-9 score of 10 or higher is considered depression and a GAD-7 score of 6 or higher is considered having symptoms of anxiety.

### Bivariate analysis of depression and anxiety in study participants

Table 3 presents the result of the bivariate analysis between dependent and independent variables, showing their relationships. Based on the results, the mean differences in depression and anxiety between women with and without functional difficulties were statistically significant (P<0.001). The mean difference in depression between pregnant and non-pregnant women was also significant (P<0.01), however, no difference in anxiety was observed between them (P>0.05). There were no mean differences in both depression and anxiety between urban and rural residents, and women gave birth or had not given birth in the past year (P>0.05). However, the mean differences between age categories, caste and ethnicity, marital status, education levels, and wealth quintiles were significant for both depression and anxiety (P<0.001). Differences between different religious groups were also significant for both depression and anxiety (P<0.05).

**Table 3:** Depression and anxiety scores and bivariate analysis by demographic characteristics

| Variables | f (%) | Depression (n=7742) |  |  |  | Anxiety (n=7742) |  |  |  |
| --- | --- | --- | --- | --- | --- | --- | --- | --- | --- |
|  |  | Mean | SD | t/F-Ratio | P-value | Mean | SD | t/F-Ratio | P-value |
| **Disability** |  |  |  |  |  |  |  |  |  |
| No difficulty | 5761 (77.4) | 2.44 | 3.46 | 44.90 | **0.000** | 3.10 | 3.42 | 59.24 | **0.000** |
| Some difficulty | 1532 (20.6) | 3.54 | 4.08 |  |  | 4.38 | 4.07 |  |  |
| A lot of difficulty | 137 (1.8) | 4.18 | 4.71 |  |  | 4.80 | 4.35 |  |  |
| Can't do at all | 12 (0.2) | 2.67 | 3.09 |  |  | 2.67 | 2.61 |  |  |
| **Currently Pregnant** |  |  |  |  |  |  |  |  |  |
| No | 7157 (96.2) | 2.67 | 3.63 | -3.42 | **0.001** | 3.39 | 3.62 | -0.72 | 0.472 |
| Yes | 285 (3.8) | 3.42 | 3.99 |  |  | 3.54 | 3.79 |  |  |
| **Birth within 1 year** |  |  |  |  |  |  |  |  |  |
| No | 6868 (92.3) | 2.71 | 3.68 | 1.45 | 0.146 | 3.41 | 3.63 | 1.83 | 0.068 |
| Yes | 574 (7.7) | 2.48 | 3.32 |  |  | 3.13 | 3.53 |  |  |
| **Place of Residence** |  |  |  |  |  |  |  |  |  |
| Urban | 4016 (54.0) | 2.65 | 3.61 | -1.18 | 0.237 | 3.38 | 3.57 | -0.34 | 0.735 |
| Rural | 3426 (46.0) | 2.75 | 3.40 |  |  | 3.41 | 3.69 |  |  |
| **Age in years** |  |  |  |  |  |  |  |  |  |
| 15 - 19 | 1379 (18.5) | 2.34 | 3.37 | 9.28 | **0.000** | 2.89 | 3.26 | 17.30 | **0.000** |
| 20 - 29 | 2489 (33.4) | 2.70 | 3.61 |  |  | 3.36 | 3.57 |  |  |
| 30 - 39 | 2031 (27.3) | 2.66 | 3.61 |  |  | 3.44 | 3.65 |  |  |
| 40 - 49 | 1543 (20.7) | 3.05 | 3.97 |  |  | 3.85 | 3.90 |  |  |
| **Marital status** |  |  |  |  |  |  |  |  |  |
| Never married | 1585 (21.3) | 2.24 | 3.11 | 45.89 | **0.000** | 2.81 | 3.12 | 57.16 | **0.000** |
| Married or living together | 5619 (75.5) | 2.75 | 3.70 |  |  | 3.47 | 3.67 |  |  |
| Single (Widowed, divorced, separated) | 238 (3.2) | 4.60 | 4.93 |  |  | 5.35 | 4.69 |  |  |
| **Religion** |  |  |  |  |  |  |  |  |  |
| Hinduism | 6307 (84.7) | 2.67 | 3.64 | 3.53 | **0.014** | 3.40 | 3.63 | 3.15 | **0.024** |
| Buddhism | 413 (5.5) | 2.40 | 3.48 |  |  | 2.93 | 3.25 |  |  |
| Islam | 275 (3.7) | 2.99 | 3.98 |  |  | 3.60 | 3.91 |  |  |
| Christianity and others | 447 (6.0) | 3.11 | 3.70 |  |  | 3.63 | 3.65 |  |  |
| **Caste & Ethnicity** |  |  |  |  |  |  |  |  |  |
| Brahmin & Chhetri | 2468 (33.2) | 2.65 | 3.63 | 9.48 | **0.000** | 3.31 | 3.50 | 16.10 | **0.000** |
| Dalit | 1242 (16.7) | 3.17 | 3.99 |  |  | 3.50 | 3.76 |  |  |
| Jana jaati | 2585 (34.7) | 2.84 | 3.73 |  |  | 3.14 | 3.49 |  |  |
| Tarai caste (Madhesi, Muslim & other) | 1147 (15.4) | 2.51 | 3.45 |  |  | 3.50 | 3.76 |  |  |
| **Education** |  |  |  |  |  |  |  |  |  |
| No school education | 2032 (27.3) | 3.06 | 4.02 | 13.75 | **0.000** | 3.89 | 3.94 | 24.34 | **0.000** |
| Basic level | 2370 (31.8) | 2.74 | 3.73 |  |  | 3.43 | 3.73 |  |  |
| Secondary | 2826 (38.0) | 2.48 | 3.32 |  |  | 3.07 | 3.28 |  |  |
| Higher | 214 (2.9) | 1.95 | 2.68 |  |  | 2.57 | 2.90 |  |  |
| **Occupation** |  |  |  |  |  |  |  |  |  |
| Not working | 1790 (24.1) | 2.43 | 3.47 | 8.65 | **0.000** | 2.99 | 3.41 | 12.40 | **0.000** |
| Professional, Managerial | 456 (6.1) | 2.36 | 3.36 |  |  | 3.20 | 3.32 |  |  |
| Sales and services | 513 (6.9) | 2.42 | 3.41 |  |  | 3.25 | 3.35 |  |  |
| Manual and others | 4683 (62.9) | 2.86 | 3.76 |  |  | 3.58 | 3.74 |  |  |
| **Wealth Index** |  |  |  |  |  |  |  |  |  |
| Poorest | 2035 (27.3) | 2.92 | 3.81 | 8.67 | **0.000** | 3.55 | 3.69 | 8.20 | **0.000** |
| Poorer | 1465 (19.7) | 2.88 | 3.83 |  |  | 3.58 | 3.83 |  |  |
| Middle | 1465 (19.7) | 2.73 | 3.72 |  |  | 3.48 | 3.76 |  |  |
| Richer | 1453 (19.5) | 2.53 | 3.36 |  |  | 3.28 | 3.42 |  |  |
| Richest | 1024 (13.8) | 2.18 | 3.29 |  |  | 2.85 | 3.19 |  |  |

### Multivariate analysis detecting factors associated with depression and anxiety

We perform logistic regression by fitting all variables into a single model to generate the odds ratio (OR) and the confidence interval (95% CI). Table 4 shows the adjusted OR for factors associated with depression and anxiety. The status of women’s disability was strongly associated with depression and anxiety (P<0.001). The odds of depression and anxiety for women with disabilities were higher by 72% and 80%, respectively, compared to those without disabilities. Similarly, the chances of depression for pregnant women were also significantly higher by 41% than for nonpregnant women (OR 1.41, 95% CI 1.06 – 1.86). Marital status was found to be strongly associated with depression and anxiety (P<0.001).

**Table 4:** Regression analysis result with adjusted odds ratio

| Variables | Depression |  | Anxiety |  |
| --- | --- | --- | --- | --- |
|  | OR | 95% CI | OR | 95% CI |
| **Disability** |  |  |  |  |
| No (Ref.) |  |  |  |  |
| Yes | 1.72*** | 1.51 – 1.97 | 1.80*** | 1.56 – 2.05 |
| **Currently pregnant** |  |  |  |  |
| No (Ref.) |  |  |  |  |
| Yes | 1.41* | 1.06 – 1.86 |  |  |
| **Marital status** |  |  |  |  |
| Never married (Ref.) |  |  |  |  |
| Married or living together | 1.19 | 0.97 – 1.48 | 1.32* | 1.07 – 1.64 |
| Single (widowed, divorced, separated) | 2.32*** | 1.65 – 3.27 | 2.31*** | 1.64 – 3.26 |
| **Religion** |  |  |  |  |
| Hinduism (Ref.) |  |  |  |  |
| Buddhism | 0.89 | 0.67 – 1.17 |  |  |
| Islam | 1.46* | 1.06 – 2.02 |  |  |
| Christianity & others | 1.33* | 1.05 – 1.68 |  |  |
| **Caste & Ethnicity** |  |  |  |  |
| Brahmin & Chhetri (Ref.) |  |  |  |  |
| Tarai caste (Madhesi, Muslims & others) | 0.96 | 0.78 – 1.19 | 1.15 | 0.94 – 1.40 |
| Dalit | 1.21* | 1.02 – 1.43 | 1.36*** | 1.15 – 1.61 |
| Jana Jaati | 0.89 | 0.76 – 1.04 | 0.90 | 0.77 – 1.05 |
| **Wealth Quintiles** |  |  |  |  |
| Poorest (Ref.) |  |  |  |  |
| Poorer | 1.08 | 0.91 – 1.27 |  |  |
| Middle | 0.92 | 0.77 – 1.10 |  |  |
| Richer | 0.82* | 0.69 – 1.25 |  |  |
| Richest | 0.78* | 0.62 – 0.99 |  |  |

\*P<0.05, \*\*P<0.01, \*\*\*P<0.001

Single women (widowed, divorced and separated) had more than two times higher odds of depression (OR 2.32, 95% CI 1.65 – 3.27) and anxiety (OR 2.31, 95% CI 1.64 – 3.26) than never married women. However, married women had a higher probability of anxiety of 32% than never-married women. Religion and wealth of the participant were also associated with depression (P<0.05). Muslims and Christians had higher odds by 46% and 33%, respectively, than Hindus. The richer and richest wealth groups had lower odds of 18% and 22%, respectively, compared to the poorest wealth groups. The caste and ethnicity of the women were also associated with both depression (P<0.05) and anxiety (P<0.001). Dalits had a higher probability of depression by 21% and anxiety by 36% than Brahmins and Chhetries.

## Discussion

The prevalence of mental health disorders in Nepal is an increasing concern as the country faces a significant mental health burden. The findings of this study found the prevalence of depression at 5.9%, while anxiety affected a substantial (22.2%) population. These findings align with the Nepal National Mental Health Survey (NMHS) conducted in 2020, which reported a lifetime prevalence of mental disorders of 10%, and 4.3% of the population experiencing a current mental disorder. Among adolescents, the prevalence was 5.2%, highlighting the vulnerability of young people to these mental health issues [26].

In a study conducted in 2016, the age and gender adjusted prevalence of the hospital anxiety and depression scale (HADS) scores for anxiety (HADS-A), depression (HADS-D) and combined anxiety and depression (HADS-cAD) were reported to be 16.1%, 4.2% and 5.9%, respectively. This study highlights the persistent prevalence of anxiety and depression as significant public health concerns in Nepal [27]. Another study of pregnant women in Nepal found that 21.3% reported anxiety and 23.8% experienced depression [20]. Significantly, being from the Dalit ethnic group was independently associated with higher rates of anxiety and depression. The prevalence of anxiety and depression among mothers of children under one year old was also concerning, with 18.7% experiencing anxiety and 15.2% experiencing depression [20].

In particular, women with disabilities in Nepal were significantly more prone to both depression and anxiety, with 72% and 80% higher chances, respectively, compared to those without disabilities. This alarming finding underscores the importance of addressing the mental health needs of individuals with disabilities [28]. Pregnant women in Nepal appeared to be disproportionately affected by mental health problems compared to non-pregnant women, with 41% higher odds of experiencing such disorders. These results are consistent with those of a study conducted in Nigeria [29], where similarly, high rates of depression and anxiety were identified among pregnant women. In the Nigerian study, 7.3% of the participants reported moderate depression, while 3.9% expressed severe anxiety, highlighting the global prevalence of these mental health challenges.

Several determinants and factors were associated with mental health disorders. Marital status emerged as a significant determinant of depression, while marital status and religion were significantly related to anxiety. In addition, factors such as marital status, educational level, and income were associated with stress among pregnant women. These findings emphasize the multifaceted nature of mental health disparities among this population [29].

Additionally, analysis indicated that marital status is associated with mental health outcomes, with single women being more likely to develop depression and anxiety compared to those who had never married. However, in the UK and Greece, marriage and social support were protective factors against depressive symptoms [28]. Caste and ethnicity were associated with both depression and anxiety, with Dalits being more prone compared to Brahmins and Chhetries. Religion and wealth status were also associated with depression. Interestingly, factors such as age, religion, education and occupation did not appear as risk factors for mental health issues in Nepal, although they showed significance in the bivariate analysis. On the contrary, a study among people with physical disabilities in Nepal found that education status and occupation were associated with depressive symptoms, with illiterate and unemployed people being more likely to experience depression [30]. Lastly, it should be noted that women in Greece and better educated individuals in the UK were more likely to experience depressive symptoms [28]. This highlights the complex interplay of cultural, social, and economic factors that contribute to mental health disparities in different contexts.

### Strengths and limitations

The present study possesses several strengths that contribute to its importance in understanding mental health in Nepal. Unlike many previous studies that were confined to specific locations and involved small sample sizes, this study benefits from being nationally representative and conducted on a large scale. The use of a nationally representative sample improves the generalizability of the findings, providing a comprehensive overview of the mental health landscape in Nepal.

However, certain limitations should be acknowledged when interpreting the findings. Firstly, the study was not primarily designed to investigate mental health and disability. Consequently, the analysis may not have captured all closely linked factors and variables that could influence mental health outcomes. Furthermore, due to its cross-sectional design, the study cannot establish causal relationships between the identified factors and depression and anxiety. Additionally, it should be noted that the study was conducted during the COVID-19 pandemic, which could have introduced unique stressors and challenges that affect mental health outcomes that may not be fully accounted for. Lastly, although the research focused on pregnant and recently delivered women, it was not specifically designed to detect perinatal depression, which may require a more specialized evaluation. These limitations underscore the need for further research to provide a more detailed understanding of mental health in Nepal.

## Conclusion

This study underscores the significant mental health challenges among women of reproductive-age in Nepal during pregnancy and delivery, with a focus on women with disabilities. The prevalence of depression and anxiety is noteworthy, affecting 5.9% and 22.2% of women, respectively. Women with disabilities are at a significantly higher risk, being 1.72 times more likely to experience depression and 1.8 times more likely to experience anxiety. Sociocultural factors such as marital status, caste, and ethnicity strongly influence mental health outcomes. Additionally, pregnancy, religion, and household wealth are associated with depression. Although the findings provide valuable information, there are limitations, including the cross-sectional nature of the data and potential pandemic-related effects. Urgent and targeted interventions and policies are needed to address mental health disparities and promote the well-being of reproductive-age women in Nepal, with a particular emphasis on those with disabilities.

## Data Availability

The study analyzed publicly available data. The data sets can be found on this website: https://dhsprogram.com/data/available-datasets.cfm

## Declarations

### Consent for publication

Not applicable.

### Authors’ contributions

HRD, SP, PA, and JB conceived and designed the study. KL extracted and HRD analyzed the data. PA and JB reviewed the literature and contributed to the analysis. HRD and SP wrote the manuscript, and all other authors reviewed, provided their input, and approved the final version of the manuscript.

## Abbreviations

ANOVA: analysis of variance
DU: dwelling unit
EA: enumeration area
GAD: generalized anxiety disorder
HADS: hospital anxiety and depression scale
ICF: International Classification of Functioning
LMIC: Low and Middle Income Country
MoHP: Ministry of Health and Population
NDHS: Nepal Demographic and Health Survey
NMHS: National Mental Health Survey
OR: odds ratio
PHQ: Patient Health Questionnaire
PSU: Primary sampling unit
SD: standard deviation

## Reference

1. Chodavadia, P., Teo, I., Poremski, D., Fung, DSS, Finkelstein, EA. Prevalence and economic burden of depression and anxiety symptoms among Singaporean adults: results of a web panel for 2022. BMC Psychiatry [Internet]. 2023;23(1):1–9. 10.1186/s12888-023-04581-7 Prevalence and economic burden of depression and anxiety symptoms among Singaporean adults: results of a 2022 Web panel | BMC Psychiatry | Full Text (biomedcentral.com)

2. Turner, RJ, Beiser, M. Major depression and depressive symptoms among physically disabled people: Assessing the role of chronic stress. J Nerv Ment Dis. 1990;178(6):343. https://pubmed.ncbi.nlm.nih.gov/2140853/

3. Chevarley FM, Thierry JM, Gill CJ, Ryerson AB, Nosek MA. Health, preventive health care, and access to health care among women with disabilities in the 1994-1995 National Health Interview Survey, Supplement on Disability. Women’s health issues. 2006;16(6):297–312. DOI: 10.1016/j.whi.2006.10.002

4. Hughes R, Swedlund N, Petersen N, Nosek M. Depression and women with spinal cord injury. Top Spinal Cord Inj Reshabil. 2001;7(1):16–24. 10.1310/HPKX-D0PV-MNFV-N349

5. Cree RA, Okoro CA, Zack MM, Carbone E. Frequent Mental Stress among Adults, by Disability Status, Disability Type, and Selected Characteristics - United States, 2018. MMWR Morb Mortal Weekly Rep. 2020;69(36):1238–43. DOI: 10.15585/mmwr.mm6936a2

6. Lund, J. I., Savoy, C., Schmidt, L. A., Ferro, M. A., Saigal, S., & Van Lieshout, RJ. influence of prenatal and postnatal adversity on depression and anxiety over two decades. J affect disord. 2020;271:178–84. 10.1016/j.jad.2020.03.138

7. Cooper PJ, Murray L, Wilson A, Romaniuk H. Controlled trial of the short- and long-term effect of psychological treatment of postpartum depression. I. Impact on maternal mood. Br J Psychiatry. 2003;182(412):9. https://pubmed.ncbi.nlm.nih.gov/12724244/

8. Crotty F, Sheehan J. Prevalence and detection of postnatal depression in an Irish community sample. Ir J Psychol Med. 2004;21(117):21. DOI: 10.1017/S0790966700008533

9. Cox, JL, Murray, D, Chapman, G. A controlled study of the onset, duration, and prevalence of postnatal depression. Br J Psychiatry. 1993;163:27–31. DOI: 10.1192/bjp.163.1.27

10. Mitra M, Iezzoni LI, Zhang J, Long-Bellil LM, Smeltzer SC, Barton BA. Prevalence and risk factors for postpartum depression symptoms among women with disabilities. Maternal Child Health J. 2015;19(2):362–72. https://link.springer.com/article/10.1007/s10995-014-1518-8

11. Wang Z, Liu J, Shuai H, Cai Z, Fu X, Liu Y et al. Mapping global prevalence of depression among postpartum women. Transl. Psychiatry. 2021;11(1):1–24. https://www.nature.com/articles/s41398-021-01663-6

12. Gajaria A, Ravindran A V. Interventions for perinatal depression in low- and middle-income countries: A systematic review. Asian J Psychiatr. 2018;37:112–20. https://www.sciencedirect.com/science/article/abs/pii/S1876201818304623

13. Nielsen-Scott M, Fellmeth G, Opondo C, Alderdice F. Prevalence of perinatal anxiety in low- and middle-income countries: A systematic review and meta-analysis. J affect disord [Internet]. 2022;306(March):71–9. Available from: 10.1016/j.jad.2022.03.032

14. Gelaye B, Rondon MB, Araya R, Williams MA. Epidemiology of maternal depression, risk factors, 1. and child outcomes in low-income and middle-income countries. Lancet Psychiatry. 2016;3(10):973–82. Available from: https://www.thelancet.com/journals/lanpsy/article/PIIS2215-0366(16)30284-X/fulltext

15. Hu HQ, Zhang J, Zhao W, Tian T, Huang HQ, Wang LL. The occurrence and determinants of anxiety and depression symptoms in women of six counties/ districts in China during pregnancy. Zhonghua Yu Fang Yi Xue Za Zhi. 2017;51(1):47–52. DOI: 10.3760/cma.j.issn.0253-9624.2017.01.010

16. Nguyen, T. V., King, J., Edwards, N., & Dunne, M. P. ‘Under great anxiety’: Pregnancy experiences of Vietnamese women with physical disabilities seen through an intersectional lens. Soc Sci Med. 2021;284(114231). https://www.sciencedirect.com/science/article/abs/pii/S0277953621005633

17. Garman EC, Schneider M, Lund C. Perinatal depressive symptoms among low-income South African women at risk of depression: Trajectories and predictors. BMC pregnancy childbirth. 2019;19(1):1– 11. 10.1186/s12884-019-2355-y

18. Pampaka, D., Papatheodorou, S. I., AlSeaidan, M., Al Wotayan, R., Wright, R. J., Buring, J. E., Dockery, D. W. & Christophi, CA. Depressive symptoms and comorbid problems in pregnancy: results from a population-based study. J Psychosom Res. 2018;112:53–8. 10.1016/j.jpsychores.2018.06.011

19. Rashid A, Mohd R. Poor social support as a risk factor for antenatal depressive symptoms among women attending public antennal clinics in Penang, Malaysia. Reprod Health. 2017;14(1):1–8. 10.1186/s12978-017-0404-4

20. Aryal KK, Alvik A, Thapa N, Mehata S, Roka T, Thapa P et al. Anxiety and Depression among Pregnant Women and Mothers of Children Under One Year in Sindupalchowk District. J Nepal Health Res Counc. 2018;16(2):195–204. https://www.nepjol.info/index.php/JNHRC/article/view/20310

21. Chalise A, Shrestha G, Paudel S, Poudyal AK. Antenatal depression and its associated factors among women of Godawari Municipality, Lalitpur, Nepal: a cross-sectional study. BMJ Open [Internet]. 2022;12(11). Available from: https://bmjopen.bmj.com/content/12/11/e063513

22. Ministry of Health and Population (MOHP), New ERA and ICF. Nepal Demographic and Health Survey. Kathmandu, Nepal; 2022. https://dhsprogram.com/pubs/pdf/FR379/FR379.pdf

23. Washington Group on Disability Statistics. The Washington Group Short Set on Functioning (WG-SS). 2017;(October):1–9. Available from: http://www.washingtongroup-disability.com/.

24. Kroenke, K., Spitzer, RL, Williams, JBW. The PHQ-9: Validity of a brief measure of severity of depression. J Gen Intern Med. 2001;16(9):606–13. 10.1046/j.1525-1497.2001.016009606.x

25. Spitzer, R.L., Kroenke, K., Williams, J.B.W., Löwe, B. A Brief Measure for Assessing Generalized Anxiety Disorder: GAD-7. Arch Intern Med. 2006;166(10):1092–7. doi:10.1001/archinte.166.10.1092

26. Nepal Health Research Council. Report of the National Mental Health Service, Nepal 2020. Vol. 5, Government of Nepal, Nepal Health Research Council. 2020. https://nhrc.gov.np/wp-content/uploads/2022/10/National-Mental-Health-Survey-Report2020.pdf

27. Risal A, Manandhar K, Linde M, Steiner TJ, Holen A. Anxiety and depression in Nepal: Prevalence, comorbidity, and associations. BMC Psychiatry [Internet]. 2016;16(1):1–9. Available from: 10.1186/s12888-016-0810-0

28. Rotarou, ES, Sakellariou, D. Depressive symptoms in people with disabilities; secondary analysis of cross-sectional data from the United Kingdom and Greece. Disabilities Health J [Internet]. 2018;11(3):367–73. Available from: 10.1016/j.dhjo.2017.12.001

29. Wegbom AI, Edet CK, Ogba AA, Osaro BO, Harry AM, Pepple BG, et al. Determinants of Depression, Anxiety, and Stress among Pregnant Women Attending Tertiary Hospitals in Urban Centers, Nigeria. Women. 2023;3(1):41–52. 10.3390/women3010003

30. Karki P, Shahi PV, Sapkota KP, Bhandari R, Adhikari N, Shrestha B. Depressive symptoms and associated factors among persons with physical disabilities in disability care homes of Kathmandu district, Nepal: A mixed method study. Public Health PLOS Glob [Internet]. 2023;3(1):e0001461. Available from: 10.1371/journal.pgph.0001461

